# Accurate SARS-CoV-2 seroprevalence surveys require robust multi-antigen assays

**DOI:** 10.1101/2020.09.09.20191122

**Authors:** Christos Fotis, Nikolaos Meimetis, Nikos Tsolakos, Marianna Politou, Karolina Akinosoglou, Vicky Pliaka, Angeliki Minia, Evangelos Terpos, Ioannis P. Trougakos, Andreas Mentis, Markos Marangos, George Panayiotakopoulos, Meletios A. Dimopoulos, Charalampos Gogos, Alexandros Spyridonidis, Leonidas G. Alexopoulos

## Abstract

There is a plethora of severe acute respiratory syndrome-coronavirus-2 (SARS-CoV-2) serological tests based either on nucleocapsid phosphoprotein (N), S1-subunit of spike glycoprotein (S1) or receptor binding domain (RBD). Although these single-antigen based tests demonstrate high clinical performance, there is growing evidence regarding their limitations in epidemiological serosurveys. To address this, we developed a Luminex-based multiplex immunoassay that detects total antibodies (IgG/IgM/IgA) against the N, S1 and RBD antigens and used it to compare antibody responses in 1,225 blood donors across Greece. Seroprevalence based on single-antigen readouts was strongly influenced by both the antigen type and cut-off value and ranged widely [0.8% (95% CI, 0.4-1.5%)-7.5% (95% CI, 6.0-8.9%)]. A multi-antigen approach requiring partial agreement between RBD and N or S1 readouts (RBD&N|S1 rule) was less affected by cut-off selection, resulting in robust seroprevalence estimation [0.6% (95% CI, 0.3-1.1%)-1.2% (95% CI, 0.7-2.0%)] and accurate identification of seroconverted individuals.

## Introduction

There is an urgent need for reliable and highly accurate SARS-CoV-2 serological tests for the diagnosis of recent or prior infection and estimation of population-wide seroprevalence (1, 2). More than 300 new SARS-CoV-2 serological tests are currently in development (updated at https://www.finddx.org/covid-19/pipeline). Assessing antibody presence with a single-readout assay is performed by selecting a cut-off value, above which the antibody is considered present (typically, three standard deviations (SD) above the negative mean distribution) (3). The majority of developed assays report high sensitivity and specificity in clinical samples obtained either from hospitalized SARS-CoV-2-infected patients or before the COVID-19 era (4). However, there is growing evidence that assays with seemingly good clinical performance might not lead to reliable diagnostic outcomes in low seroprevalence epidemiological studies, where asymptomatic carriers with low antibody titers are overrepresented (2, 5). One approach that holds promise in addressing this issue, is the development of multi-antigen assays coupled with consensus-based rules.

Recently, a small number of multi-antigen SARS-CoV-2 serological assays that employ “AND” and “OR” logic rules between different antigen readouts have been developed, showing improved performance in clinical samples (6-9). More specifically, due to their consensus-based approach, multi-antigen assays report increased specificity, without the cost of reduced sensitivity, compared to their single-antigen counterparts. Furthermore, because of the multiple antigen readouts, they can increase the certainty of diagnosis in borderline cases, i.e. low-titer positive samples (10). To the best of our knowledge, how this enhanced clinical performance and robustness of multi-antigen approaches translates to low seroprevalence epidemiological studies has yet to be experimentally investigated.

In this study, we developed a bead-based multiplex serological assay for the simultaneous detection of antibody response against the N, S1 and RBD SARS-CoV-2 antigens and applied it to a low seroprevalence epidemiological study. The multi-antigen approach requiring partial agreement between single-antigens (RBD&N|S1 rule) was first evaluated in well-defined clinical samples and validated against commercially available single-antigen assays, showing improved performance. In order to experimentally elucidate how this increase in performance transfers to an epidemiological setting, we compared antibody responses against the N, S1 and RBD antigens in 1255 blood donors across Greece (26% female; median age, 42 years; range, 17-65). We report striking differences in seroconversion calls between the different antigens that resulted in a wide range of estimated seroprevalence rates and conflicting diagnoses of positive individuals. On the other hand, the multi-antigen RBD&N|S1 approach resulted in a more robust estimate of seroprevalence and accurate identification of seroconverted individuals.

## Results

### Clinical performance of SARS-CoV-2 multiplex assay

The clinical performance of the developed multiplex assay was evaluated in 155 serum samples from 77 PCR-confirmed COVID-19 cases and 78 pre-epidemic individuals. The sensitivity and specificity of the assay were calculated for the three single antigen readouts and for 11 multi-antigen consensus-based rules for detection of total, IgG, IgA and IgM isotypes. Performance was evaluated using a cut-off value of mean plus three standard deviation (SD) for each antigen and the results are presented in Table 1. When assessed individually, antigens were equally specific producing one or two false positives out of the 78 negative samples tested [specificity 97.4% (95% CI, 91.1-99.3%) to 98.7% (95% CI, 92.7-99.8%)]. Rules-based approaches, requiring at least two of the antigens to be above the cut-off (i.e. rules using an AND gate between two antigens) for reporting a positive result, improved assay specificity in all isotypes *vs*. individual antigens (Table 1). On the other hand, rules, requiring at least one/any of the antigens to be above the cut-off (i.e. OR-based rules), did not improve assay performance when compared to the best performing individual antigen for the respective antibody isotype. Overall, rules that utilized all three antigens and set a positive result when at least two of them were above the cut-off (Antigen A AND [B OR C] reported as A&B|C) showed improved and consistent performance and were further investigated. Similar results were obtained when different cut-off values, based on ROC analysis, were used (see Supplementary Table S6 and Supplementary Figure S4). A detailed analysis of antibody response for the clinical samples is presented in Supplementary Material S1.3.

**Table 1.**
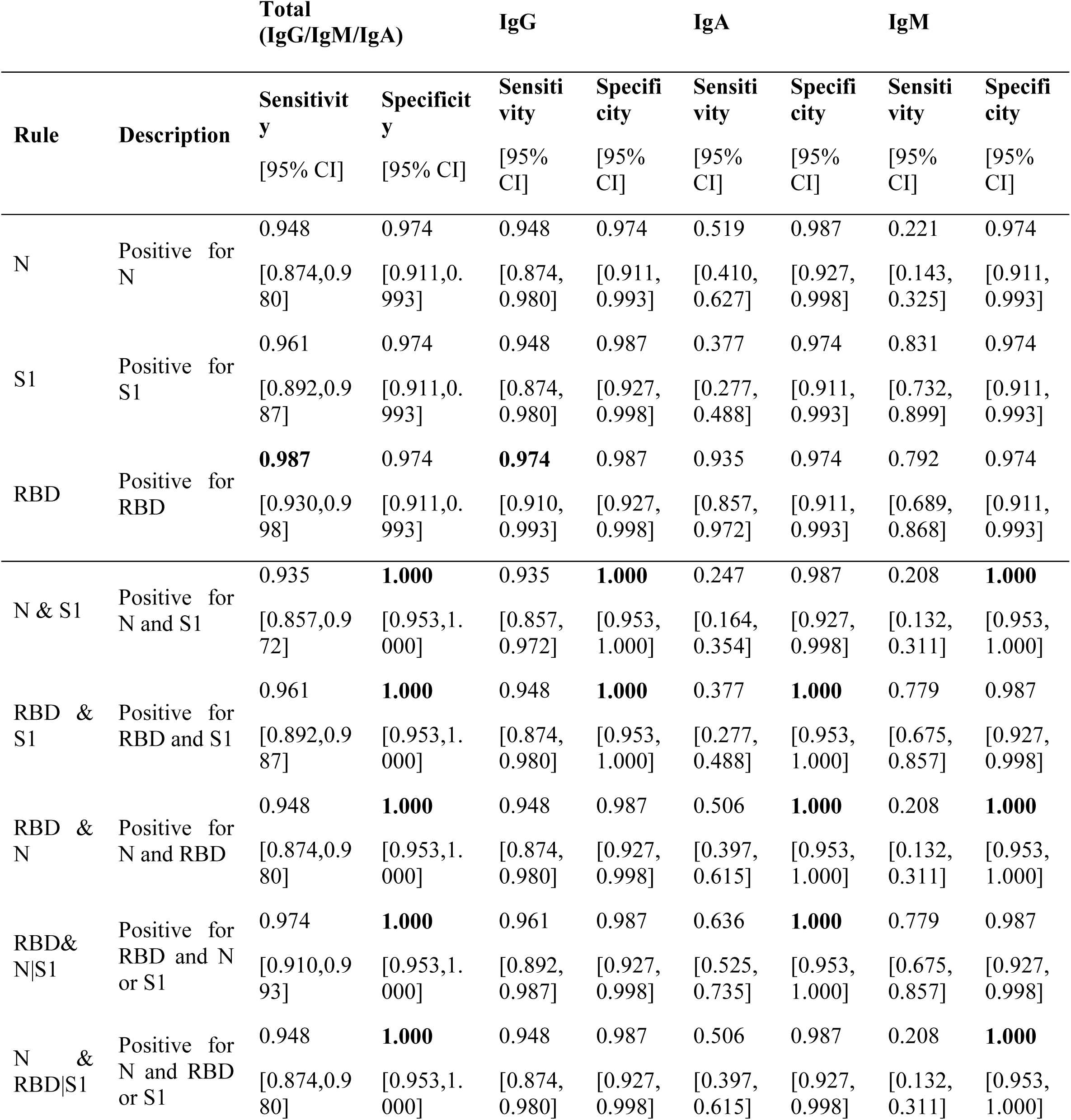
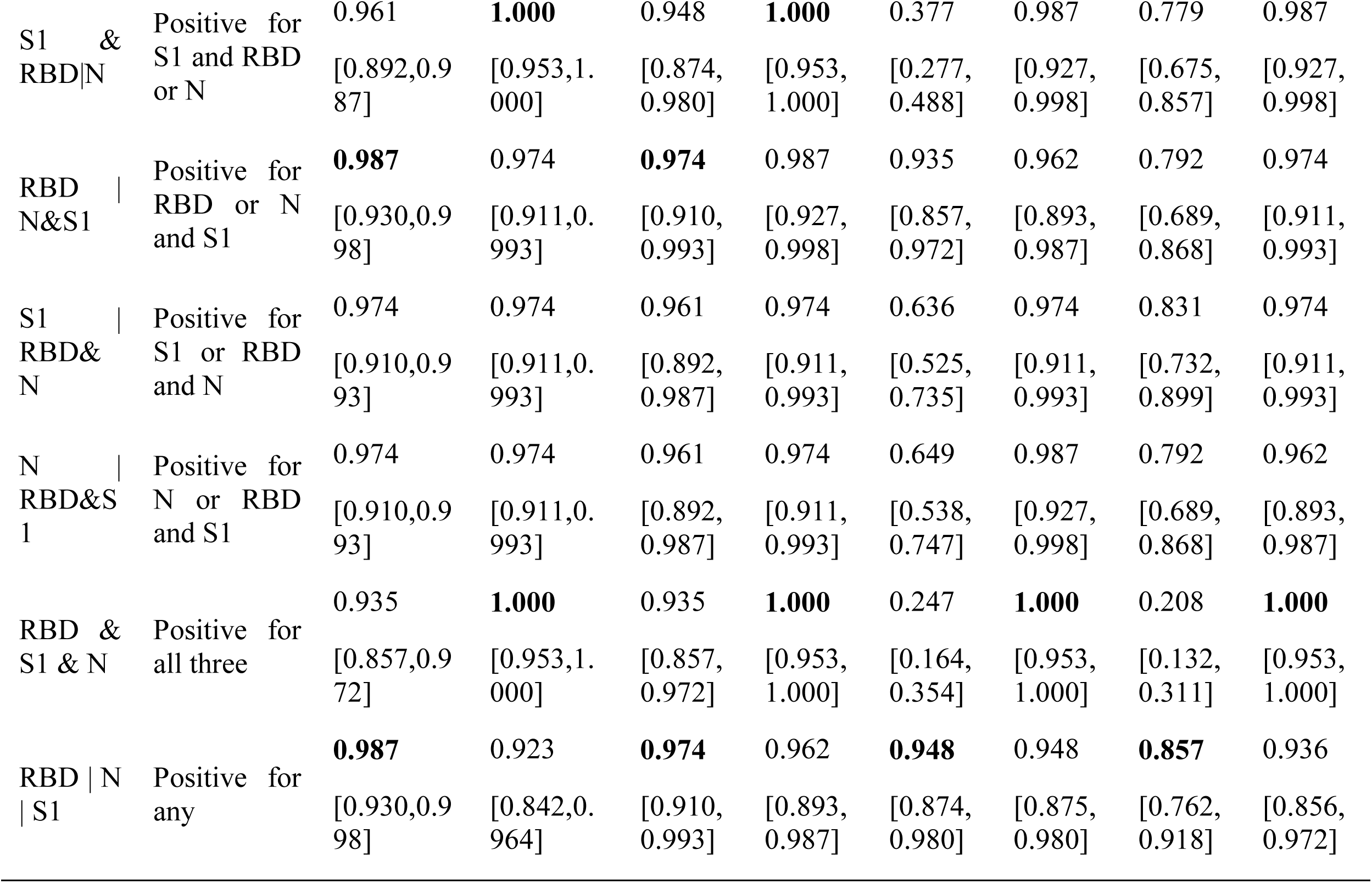
Clinical sensitivity and specificity of single-antigen assays and multi-antigen rules.

To assess the robustness of the multi-antigen rules their performance was evaluated for a wide range of cut-off values. Sensitivity, specificity and accuracy were calculated for gradually increasing threshold (cut-off) values, based on the negative sample distribution of each antigen and antibody isotype and are presented in Figure 1. Rules using all antigens and requiring two or all three to be higher than the threshold were included in the analysis. Across all isotypes, rules exhibited a more robust profile and were less affected by changes in the cut-off thresholds compared to individual antigens. Rules provided a clear benefit in assay specificity with specificities of 100% (95% CI, 95.3-100%) achieved at much lower cut-offs compared to individual antigens. Additionally, for total and IgG antibody detection, assay sensitivity was retained at high levels across a wide range of cut-offs resulting in an overall more robust and accurate assay. The RBD&N|S1 rule was shown to outperform all other rules in terms of robustness and performance (Figure 1).

**Figure 1.**
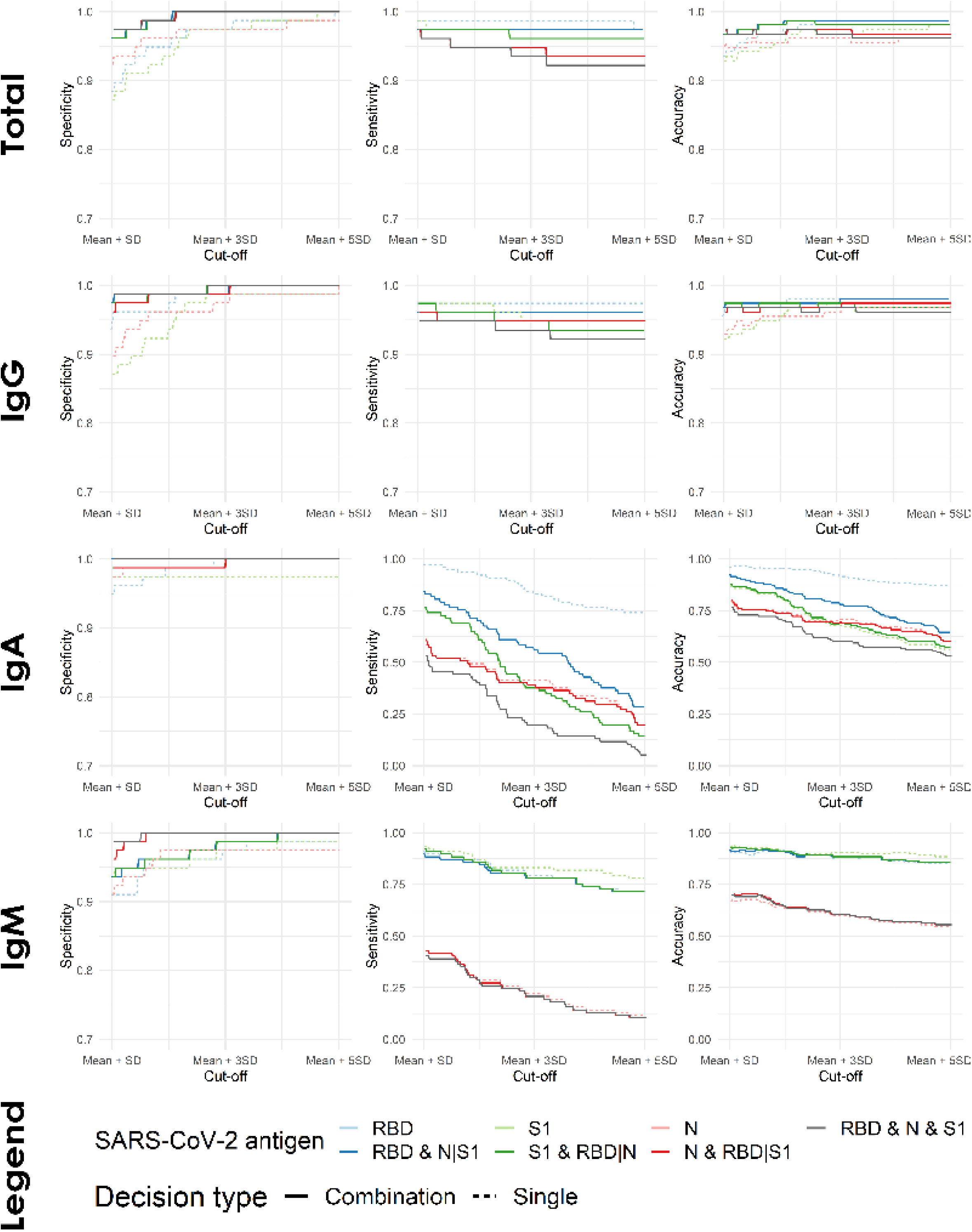
Assay performance metrics for different cut-off thresholds. For each antibody isotype, sensitivity, specificity, and accuracy values were calculated for increasing cut-off thresholds based on the distribution of each antigen’s negative samples.

### Assay validation against commercial antibody tests

The performance of our assay was validated using commercially available antibody tests developed by Abbott and Euroimmun AG against the N and S1 antigens, respectively (Figure 2). As shown in Figure 2A, both N and S1 antigen readouts of the multiplex assay were highly correlated with their commercial counterparts (Pearson’s r=0.98 for N and 0.9 for S1). Their diagnostic agreement (positive/negative call) was 100% when compared to Euroimmun S1 assay and Abbott N assay. A strong agreement was also observed between the commercial assays and the RBD&N|S1 multi-antigen rule (Figure 2B). Specifically, the RBD&N|S1 rule agreed in 59 out of the 60 samples tested with the Euroimmun S1 assay and in 29 out of the 31 samples tested with the Abbott N assay. Finally, the three samples in which the assays disagreed, were called negative by the commercial assays but showed positive readouts in the other two antigens measured in our multiplex assay and were thus called positive by the multi-antigen rule.

**Figure 2.**
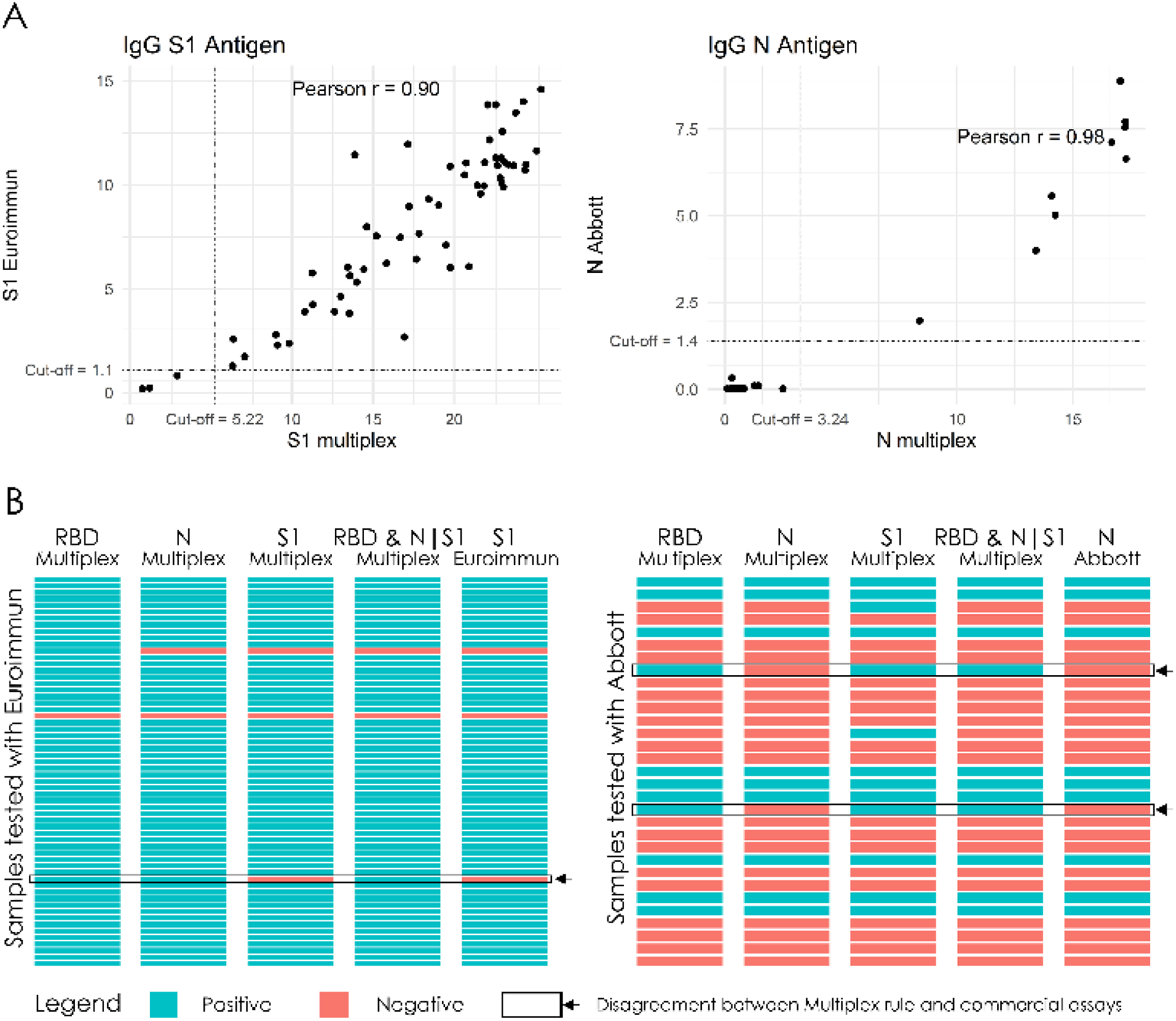
Validation of the multiplex assay against commercial serological assays. (A) Scatter plot of the S1 and N single-antigen readouts of the multiplex IgG assay (S1 multiplex and N multiplex) against results from the Euroimmun S1 (n=60) and Abbott N (n=31) commercial assays, respectively. Dotted lines correspond to assay cut-offs for positivity. (B) Heatmap of diagnostic outcomes depicting the agreement between the multiplex and the commercial assays.

### Seroprevalence survey

An important application of SARS-CoV-2 serological assays is the identification of seroconverted individuals at the population level. Given that asymptomatic infection may induce low SARS-CoV-2 related antibodies titers which may also decline over time, a major requirement for this type of analysis is a highly sensitive and specific assay. Therefore, we used the total (IgG/IgA/IgM) SARS-CoV-2 antibody multiplex assay to assess how its high clinical performance translates to estimating seroprevalence in 1,225 asymptomatic blood donors with no known history for SARS-CoV-2 exposure. Seroprevalence was calculated based on the single antigen readouts and the multi-antigen RBD&N|S1 rule and was found to be strongly influenced both by the antigen analyzed and the cut-off value used to determine the diagnostic outcome (Figure 3A). As it can be seen in figure 3A, seroprevalence rates from single antigen readouts ranged between 0.8% (N, mean plus 5 SD cut-off, 95% CI 0.4-1.5%) and 7.5% (S1, mean plus 3 SD cut-off, 95% CI 6.0-8.9%), indicating a wide range of potentially indeterminate cases. When using the RBD&N|S1 rule, seroprevalence ranged between 0.6% (mean plus 5 SD cut-off for each antigen, 95% CI 0.3-1.1%) and 1.2% (mean plus 3 SD cut-off, 95% CI 0.7-2.0%), in line with the robust performance of the multi-antigen assay in clinical samples. Furthermore, we examined the overlap of positive individuals being diagnosed by the single antigens or the RBD&N|S1 rule using the stringent cut-off of mean plus 5 SD (Figure 3B). We observed a strikingly low agreement between antigens, in that different antigens resulted in vastly different subsets of positive individuals. Specifically, N shared 2 positive samples with RBD (5.9% agreement) and 1 with S1 (1.8% agreement), while S1 and RBD shared 6 positive samples (9.1% agreement). The RBD&N|S1 rule had a total of 7 positive calls, 5 of which were samples with S1 and RBD positive readouts, 1 with RBD and N positive readouts and 1 with all three antigens above the cut-off. Finally, we re-analyzed all 1,225 samples with an independent commercially available test which detects IgG N-specific antibodies (Abbott). From the 6 samples that were called positive (estimated seroprevalence 0.5%) only 2 also scored positive with the multiplex RBD&N|S1 rule. An analysis of the seroprevalence, agreement rates and overlap of positive samples between the other rules of the multiplex assay (N&RBD|S1, S1&RBD|N) and Abbott’s test results (N-specific IgG) are shown in Supplementary Figures S5, S6 and S7.

**Figure 3.**
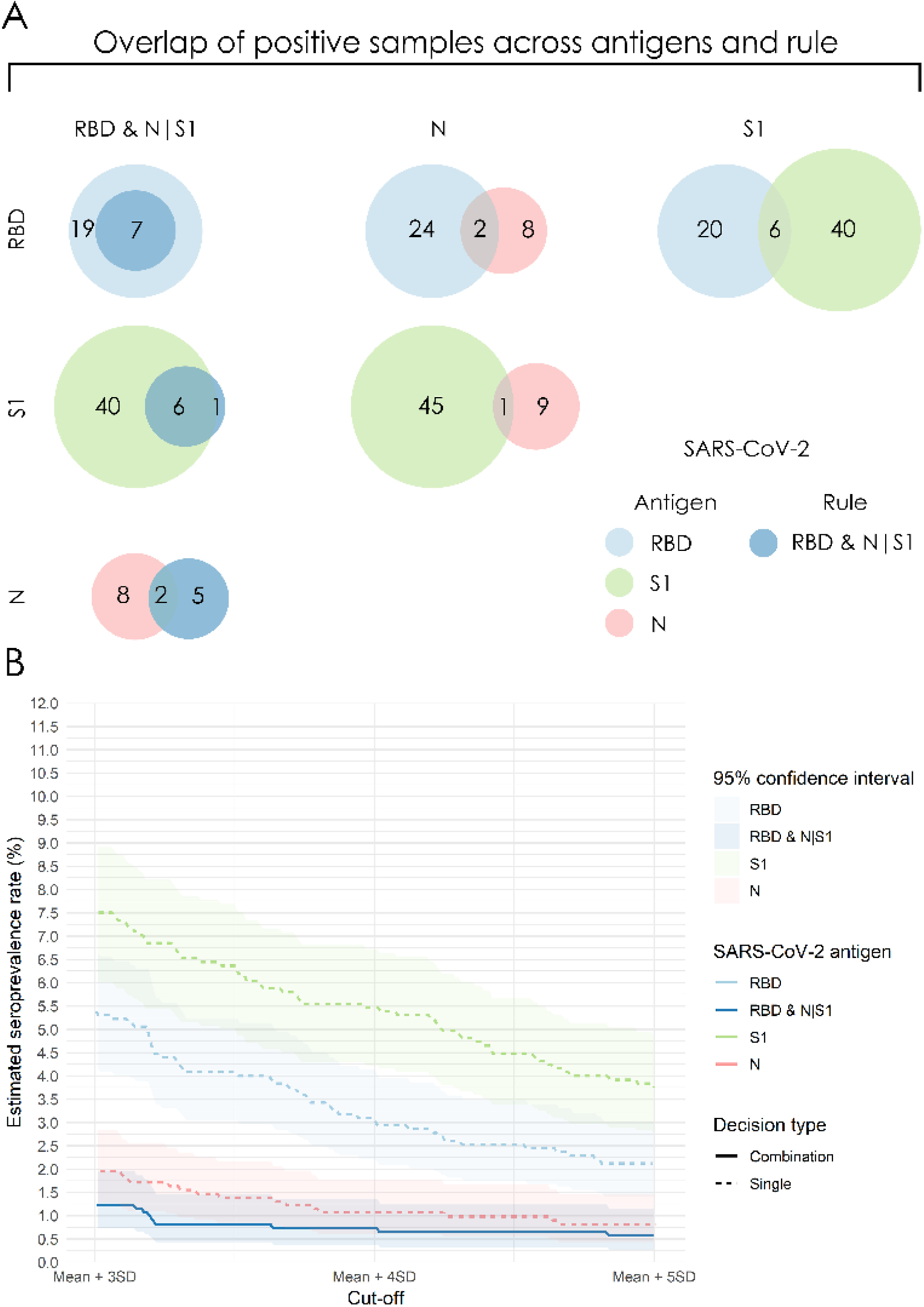
SARS-CoV-2 seroprevalence analysis in 1,225 blood donors using single-antigen readouts or the RBD AND (N OR S1) multi-antigen rule (RBD&N|S1). (A) SARS-CoV-2 estimated seroprevalence rates using single-antigen readouts or the RBD&N|S1 rule for different cut-off values (RBD: Receptor Binding Domain, N: nucleocapsid phosphoprotein, S1: S1 subunit of spike glycoprotein, SD: standard deviation above the negative mean). (B) Consensus in detection of SARS-CoV-2 positive individuals using single and multiple-antigen readouts.

## Discussion

During the COVID-19 pandemic, serological tests play an instrumental role in quantifying seroconversion, seroprevalence, vaccination status and diagnosing recent or prior SARS-COV2 infection. More than 300 assays have been developed using either the N, S1 or RBD antigens and many of them have already reached the market in unforeseen development speeds (4). However, it is questionable whether single-antigen assays, with strong clinical performance characteristics, show sufficient diagnostic performance in population screening studies (5). Here, we developed single and multi-antigen assays, evaluated their performance in clinical samples and investigated how the increased performance of multi-antigen assays translates to seroprevalence estimation in a population study.

The developed multiplex platform enabled the simultaneous detection of diverse antibody responses against N, S1 and RBD, within a single diagnostic run. In line with other clinical performance reports, we found a high variability of IgM and IgA responses to different antigens in 77 previously infected SARS-CoV-2 individuals (Supplementary Figure S3) (11). In contrast, nearly all positive individuals produced IgG specific antibodies against all three antigens. Other studies have also evaluated N, S1, and RBD antigens in single or multiplex formats and reported similar performance characteristics in samples tested two to three weeks post infection (6, 12-14). The validation of our single-antigen assays against commercial single-antigen tests (Abbott-N and Euroimmun-S1) showed 100% agreement with our N and S1 readouts (Figure 2). However, diagnostic outcomes between commercial tests and the RBD&N|S1 rule were slightly different, suggesting that previously infected SARS-CoV-2 individuals may be misdiagnosed by single antigen-based tests (Figure 2B). In support, the absence of antibodies against certain SARS-CoV-2 antigens is also apparent in studies that have compared assays targeting different antigens (6, 14-16); yet, the biological basis of these observations is currently not well understood.

The rules-based approach combines single antigen readouts, using “AND” and “OR” logic rules, in order to further enhance the specificity, accuracy and robustness of the multi-antigen assay. Such approaches have already been shown to enhance the diagnostic performance of serological assays for common SARS-CoV (17) and for SARS-CoV-2 (9). In our study, we showed that an optimally selected combination of AND plus OR rules can increase specificity and accuracy, with little or no cost on sensitivity, for IgG and total antibody detection in clinical samples (Table 1). After testing all potential rule combinations the optimal rule was identified as the RBD&N|S1 (Table 1). The optimal rule utilized the best performing readout (RBD antigen) whose specificity was further enhanced by requiring consensus (AND) with either S1, N, or both readouts. Most importantly, the rules-based approach improved assay robustness (i.e. how small deviations of the cut off value affect the diagnostic outcome). The RBD&N|S1 rule resulted in an assay with robust performance across a wide range of cut-off values, compared to single-antigen assays, suggesting that this multi-antigen combinatorial assay can achieve optimal performance at lower cut-offs (Figure 1). It is worth noting that while multi-antigen rules exhibited improved performance in IgG and total antibody detection, they did not improve the performance of the IgA and IgM assays (Table 1).

The use and application of our multiplex strategy in 1255 samples from randomly selected blood donors aims to provide a dataset of experimental settings that mimic routine low-prevalence screening. To the best of our knowledge, this is the first time a multi-antigen serological assay was applied to a seroprevalence study. In contrast to clinical samples, where almost all positive donors showed high antibody titers for all three antigens, a substantial number of donors revealed different responses against the N, S1 and RBD antigens and only one of the 1,225 donors showed high antibody titers against all three antigens. With such high dissensus between single-antigen responses, it is obvious that diagnosis of seroconversion in the population becomes challenging. Even the highly concordant S1 and RBD antigens exhibited only 9% agreement in their positive calls (6 out of 66 positive calls) (Figure 3B). One explanation for this disagreement is that SARS-CoV-2 infected individuals in population-wide screens are mostly asymptomatic, with low antibody titers and unknown time since infection, during which antibodies to specific epitopes may have already waned below the detection limit (18-20). Another important reason for this large disagreement is the imperfect specificity of single-antigen assays, which can lead to a large number of false predictions in a population screening (5). For example, assuming 1% seroprevalence and 100% sensitivity, the maximum observed 97.4% single-antigen specificity in the clinical performance study would result in ∼31 false positive predictions in the serosurvey, whereas just 12 samples are expected to be truly positive (21). Based on this, we believe that the estimated 7.5% seroprevalence, which was observed in Figure 3A at mean plus 3 SD with S1, may include mostly false positive predictions. The striking differences in the seroconversion calls between antigens were reflected not only in the wide range of estimated seroprevalence rates (0.6%-7.5%) but more importantly in the vastly different subsets of potentially SARS-CoV-2 positive individuals (Figure 3B). Consistent to our findings, a side-by-side comparison of three fully automated SARS-CoV-2 antibody assays (Abbott against N, Roche against N, and DiaSorin against S1/S2 antigens) showed good agreement in 65 samples from COVID-19 patients, but had profound discrepancies in positive predictions at 1% seroprevalence (22). Likewise, in an epidemiological study in Iceland, out of the 18,609 individuals tested for total anti-N and anti-S1-RBD antibodies using independent commercial kits, 158 were found positive for either N or S1-RBD but only 39 of them had antibodies against both epitopes (23). Similar discrepancies between N- and S1-specific serological assays were observed in a large epidemiological study in Spain (24).

Our results have profound implications regarding seroprevalence rates presented by studies based on single antigen assays, as well as for their diagnostic value. To achieve the most accurate estimate of seroprevalence, an ideal 100% specific assay is required. One approach to increase specificity involves raising the cut-off value of the assay, an approach that IVD manufacturers prefer to adopt to be on the safe side regarding positive predictions. However, raising the cut-off value of single-antigen approaches can lead to a profound underestimation of the seroprevalence rate (as shown in Figure 3A). A better approach for accurate seroprevalence estimation would be confirmatory testing with an independent assay that uses a different antigenic target (25). In this paper, we showed that the specificity of the multi-antigen assay can be increased by using a consensus-based strategy, rather than increasing the cut-off value (Table 1). On this front, our rules-based method achieved an almost consistent seroprevalence rate in a wide range of cut-off values between 3 to 5 standard deviations above the mean (Figure 3A). Consequently, we believe that the RBD&N|S1 rule combined with a mean plus 5 SD threshold provides a realistic estimation of seroprevalence in the community and accurate identification of seroconverted individuals. Notably, in our study we used cut-off thresholds between 3 to 5 SDs whereas the manufacturer’s recommended 1.4 cut-off value for the IgG N-specific Abbott test was calculated to correspond to more than 10 SDs above the negative mean. Such high cut-off values can undermine sensitivity and result in the underestimation of the true seroprevalence rate. When using the rules-based multiplex assay (RBD&N|S1) as reference the positive predictive value of the IgG N-specific Abbot test was only 29% (2 out of 7 detected), despite both assays showing excellent agreement in clinical samples (Supplementary Figure S7). An important limitation of such comparisons between serological assays in population-wide surveys is the fact that there is no gold standard method to identify the truly asymptomatic SARS-CoV-2 positive cases.

In conclusion, our study has demonstrated that serological assays based on single antigens, while good at diagnosing infected individuals in a clinical setting, may not be ideal in low seroprevalence, population-wide COVID19 screens, where low antibody responses from mostly asymptomatic individuals are expected. A multi-antigen approach combined with a rules-based framework for diagnostic decisions can provide a better alternative in this context, through its enhanced specificity and reduced dependency on cut-off values. We believe that such multi-antigen approaches should be performed in a single multiplex assay, thus diminishing possible differences attributed to operational issues of independent assay formats (11). An added advantage of multiplexing is the reduced usage of resources and time. The embrace of multiplex assays or multiple single antigen-based assays, by the scientific community, for epidemiological studies can eventually lead to more accurate and reliable results regarding SARS-CoV-2 spread in the population.

## Methods

### Serum samples

A total of 155 clinical serum samples were analyzed, of which 78 were negative as were banked sera collected prior to the COVID-19 pandemic (2018-2019) and 77 were positive samples from PCR-confirmed SARS-CoV-2 individuals collected at least two weeks after SARS-CoV-2 infection. Nineteen additional serum samples from PCR-confirmed SARS-CoV-2 individuals were used for assay validation against other commercial SARS-CoV-2 serological assays. For the general population screening, we obtained 1,225 serum samples between the 23^rd^ and 25^th^ week of 2020, e.g. 5 weeks after lockdown in Greece that took place between the 13^th^ and 18^th^ week of 2020; blood donors were from 13 different geographical regions in Greece (26% female; median age, 42 years; range, 17-65). Eligibility for donation included an extended detailed questionnaire for previous possible signs of infection during 2020. All samples were acquired under approved clinical protocols and informed consent (see Ethics statement). The detailed list of serum samples used throughout the study is presented in Supplementary Table S2.

### Multiplex immunoassay development and clinical performance analysis

A magnetic bead-based immunoassay was developed using the xMAP Luminex technology against SARS-CoV-2 antigens N, S1 and RBD. Additionally, one antigen from each one of the four endemic coronaviruses was also included in the assay. Specifically, the S1 subunit of HCoV-HKU1, HCoV-229E and HCoV-NL63 and the S1+S2 subunits from HCoV-OC43 were used. For details regarding the optimization parameters and technical specifications of the multiplex assay see Supplementary Material S1.1. The Median Fluorescent Intensity (MFI) values of each SARS-CoV-2 antigen were first divided by the average MFI of the negative control samples (made from a pool of negative sera) for the same antigen. Cut-off values for determining the diagnostic outcome (positive/negative) regarding the presence (or not) of SARS-CoV-2 specific antibodies were calculated for each SARS-CoV-2 antigen based on its negative sample distribution. Performance was evaluated in terms of sensitivity, specificity and accuracy, while the corresponding 95% confidence intervals (CI) were calculated using the Wilson approximation (26). The Receiver Operating Characteristic (ROC) analysis of the assay is presented in Supplementary Material S1.5.

### Rules-based method

A rules-based method was developed to combine the readouts of the multi-antigen assay. First, the single antigen “normalized MFI” values were transformed to positive/negative predictions by comparing them with the appropriate cut-off value. Then, logic circuits that utilize the “AND” (represented by the symbol “&”) and “OR” (represented by the symbol “|”) logic gates were implemented. These logic circuits take as input the single-antigen predictions and output the final rules-based prediction that corresponds to a positive/negative call for the presence of SARS-CoV-2 specific antibodies. All possible simple circuits that could be formed using the “AND” and “OR” logic gates to combine the predictions of the RBD, S1 and N predictions were examined.

### Assay validation

Assay validation was performed in two separate subsets of matched clinical samples against two widely used, commercially available SARS-CoV-2 antibody tests developed by Euroimmun (Euroimmun Medizinische Labordiagnostika AG, Lubeck, Germany) and Abbott (Abbott Diagnostics, Illinois, USA), which detect IgG antibodies against S1 and N respectively (Supplementary Table S7). Single antigen readouts for S1 and N from our IgG multiplex assay were plotted against the results of the commercial tests and the Pearson’s correlation coefficient was calculated. For determining the diagnostic outcome (positive/negative calls) we used the mean plus five SD cut-off values for our assays and the manufacturer’s recommended cut-offs for the commercial assays (1.1 for Euroimmun and 1.4 for Abbott).

### Population-level analysis

For the analysis of the 1,225 samples from blood donors, the total (IgG/IgA/IgM) assay was used, and the diagnostic outcome was assessed across cut-off values ranging from mean plus 3 SD to mean plus 5 SD from the negative sample distribution. To identify seroconverted individuals, the diagnostic outcomes using the mean plus 5 SD cut off values were utilized. Frozen back-up samples (n=1,225) were sent to the Immunology Laboratory of the National Public Health Organization, Athens, Greece and analyzed for the presence of IgG antibodies against the N antigen using the Abbott IgG assay with the ARCHITECT i2000SR analyzer (Abbott, Illinois, United States). The manufacturer’s recommended cut-off (1.4) was used to determine positivity.

## Data Availability

The datasets generated during and/or analysed during the current study are available from the corresponding author on reasonable request.

## SUPPLEMENTARY MATERIALS

**Supplementary Results 1:** 1) Optimization of assay Parameters. 2) Analytical assay performance. 3) Antibody responses to SARS-CoV-2 and endemic human coronaviruses in clinical samples 4) Correlation of SARS-CoV-2 antibody responses to antibody levels from other infectious agents 5) Receiver operating characteristic curve analysis. 6) Seroprevalence and agreement rates of single and multi-antigen rules in the population screening 7) Assay Validation. 8) Seroprevalence and agreement rates of single and multi-antigen rules in population screening.

**Supplementary Methods 2:** 1) Multiplex immunoassay development 2) Clinical and donor samples 3) Assay validation

**Figure S1.** Serum Dilution Optimization.

**Figure S2.** Secondary Antibody Dilution Optimization.

**Table S1.** Intra- and Inter-assay variability, measured as % coefficient of variation.

**Table S2.** List of samples used throughout this study.

**Figure S3.** Antibody responses against SARS-CoV-2 N, S1 and RBD antigens and 4 endemic coronaviruses antigens in serum samples from SARS-CoV-2 PCR-tested positive and negative (banked blood samples from 2018-2019) individuals.

**Table S3.** Antibody responses to coronavirus antigens in SARS-CoV-2 positive and negative cases.

**Table S4.** Correlation of normalized MFI values between SARS-CoV-2 antigens for the different antibody isotypes in positive and negative cases.

**Table S5.** Correlation of SARS-CoV-2 antibody responses to antibody levels from other infectious agents.

**Figure S4.** ROC curves for the 3 antigens and different antibody isotypes. Confidence intervals are demonstrated with red dotted lines.

**Table S6.** Diagnostic accuracy and AUC of SARS-CoV-2 individual antigens.

**Figure S5.** Percentage of asymptomatic individuals (n=1,255) positive for total (IgG/IgA/IgM) SARS-CoV-2 antibodies using single antigen readouts and multi-antigen rules.

**Figure S6.** Consensus between positive samples for total (IgG/IgA/IgM) antibodies using N, S1, RBD and respective multi-antigen rules in the population screen.

**Figure S7.** Consensus between positive samples for total (IgG/IgA/IgM) antibodies against N, S1 and RBD and for IgG antibodies against N antigen (Abbott) in the population screen.

**Table S7.** Commercial SARS-CoV-2 serological assays and samples used for assay validation.

## ACKNOWLEDGMENTS

We thank the patients and individuals who donated their blood. We greatly acknowledge all healthcare workers who were involved in this study.

## AUTHOR CONTRIBUTIONS

Conceptualization: LGA, AS; Cohort management and sample collection: MP, KA, ET, AM, MM, GP, MAD, CG, AS; Methodology and sample measurements: CF, NM, NT, VP, AM, AM; Data Analysis: CF, NM, NT, IPT, AS, LG; Writing: CF, NM, NT, IPT, AS, LG; All authors have read and agreed to the final version of the manuscript.

## ETHICAL STANDARDS

Sampling from SARS-CoV-2 positive individuals was done with informed consent and under approved institutional and ethic review board approved clinical protocols conducted in full compliance with the principles of Good Clinical Practice and the Declaration of Helsinki (University Hospital of Patras EC 164/27.04.2020 and IRB 216/08.05.2020, Alexandra General Hospital NCT04408209 trial). Stored negative samples were acquired in accordance with local ethical approvals. The population-level study was approved by the Research Ethics Committee of the University of Patras (Ref. Number 6099) and all participants gave written informed consent.

