## Supplemental Material for "Accurate SARS-CoV-2 seroprevalence surveys require robust multi-antigen assays"

[#]co-principal investigators

**Corresponding author:** Leonidas G. Alexopoulos

### Supplementary results

#### S1.1 Optimization of assay parameters

Serum dilution and antibody detection concentration were optimized during assay development using a small subset of well-defined positive samples for prior SARS-CoV-2 infection and negative serum samples (Supplementary Figures S1 & S2). Samples were assessed at four two-fold serial dilutions (1:100, 1:200, 1:400 and 1:800). A serum dilution of 1:400 was found to provide the best ratio of positive to negative MFI values in total (IgG/IgM/IgA), IgG and IgM isotype detection and was chosen for subsequent experiments. For IgA detection, the optimal serum dilution was 1:100. The biotinylated detection antibodies were also tested at two-fold serial dilutions from 1:800 to 1:6,400 and the signal-to-noise ratio (SnR) of each serum sample to a blank sample was calculated. The antibody concentrations that increased SnR values in positive samples and/or decreased SnR values in negative samples were considered optimal. A dilution of 1:800 was chosen for IgG and IgM isotypes, while antibodies against total and IgA isotypes were used at 1:1,600 and 1:3,200 dilution, respectively.

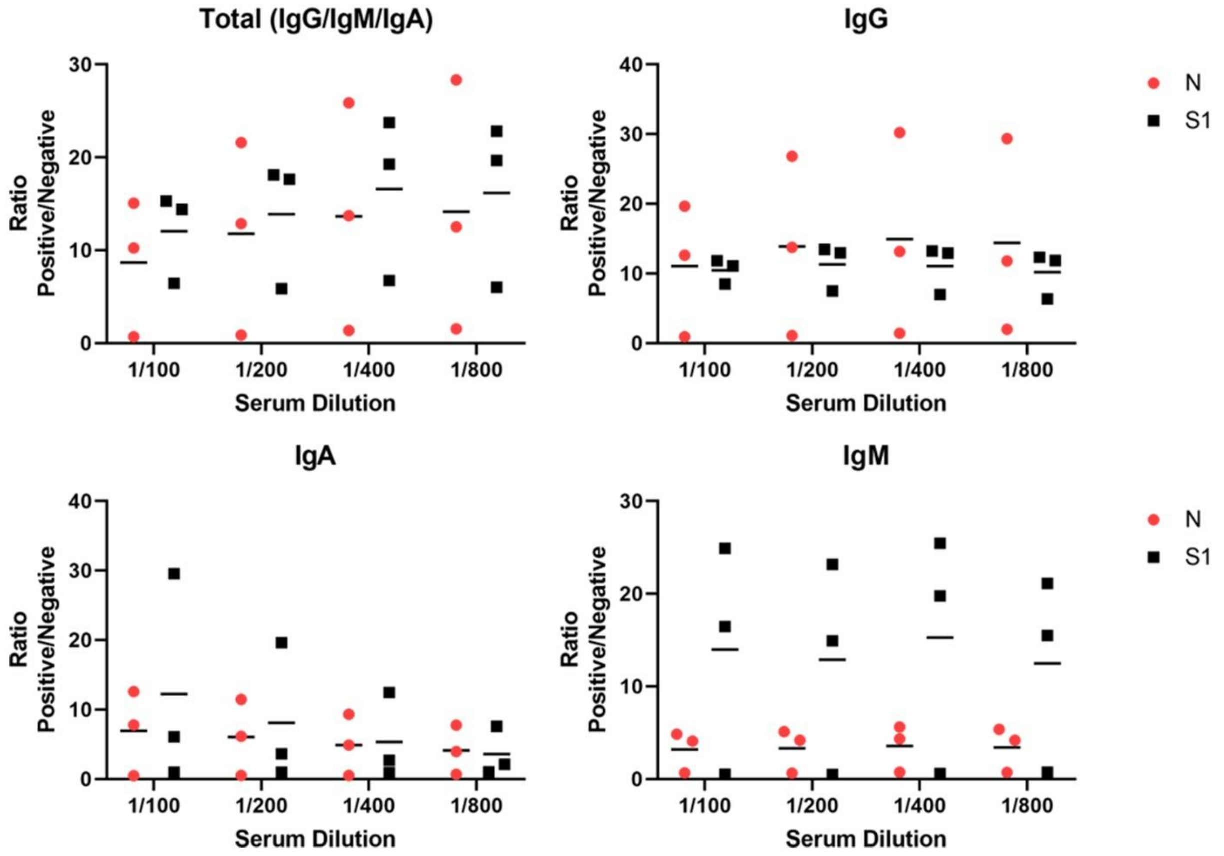


**Supplementary Figure S1.** Serum Dilution Optimization. Serum from three SARS-CoV-2 positive samples and four negative samples cases were tested at four dilutions and the ratio of the MFI in each positive case to the average MFI from the negative cases was calculated and plotted for N and S1 antigens. Black lines represent the mean of the three positive samples.

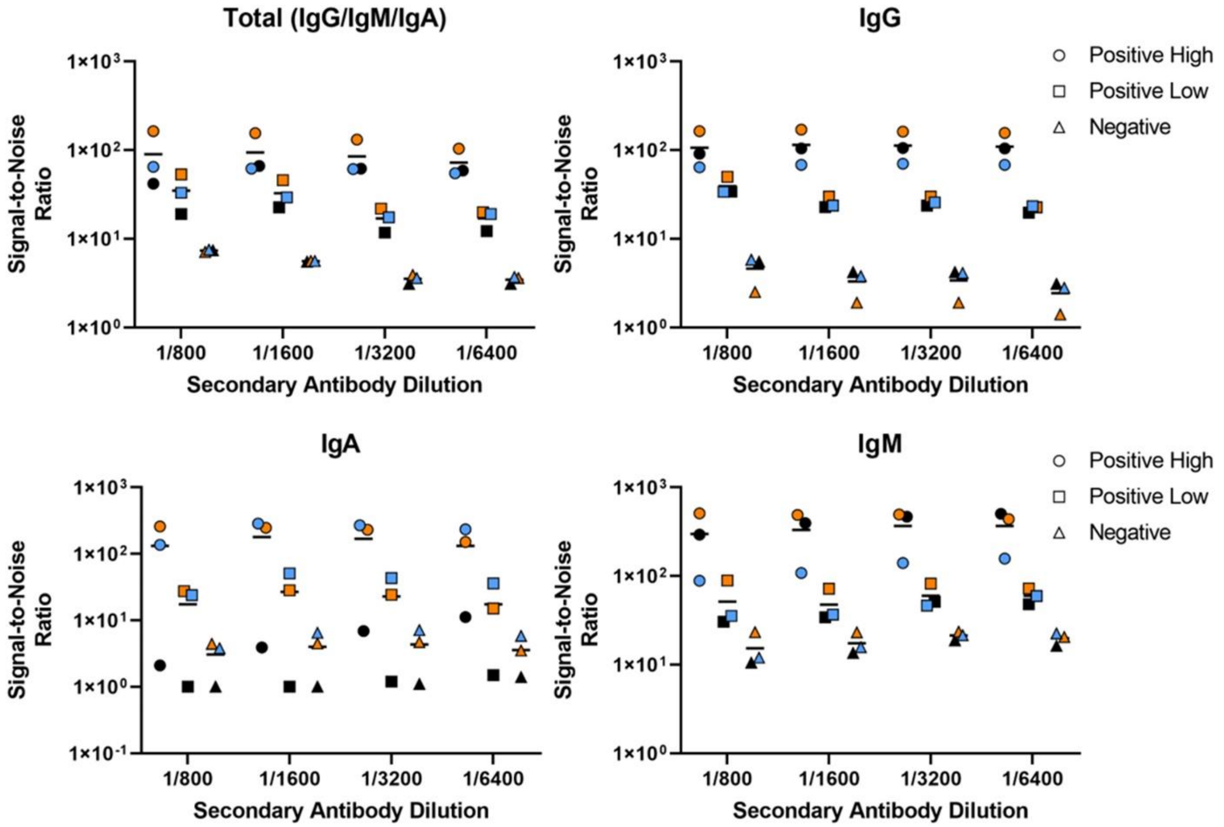


**Supplementary Figure S2.** Secondary Antibody Dilution Optimization. A SARS-CoV-2 positive serum with high antibody levels (positive high - circles), a SARS-CoV-2 positive serum with low antibody levels (positive low - squares), a negative serum (triangles) and a blank sample (serum diluent only) were tested at four two-fold serial dilutions of the secondary antibody. For each positive and negative sample, the signal-to-noise ratio against the blank sample was calculated for N (black), S1 (orange) and RBD (blue) and plotted against the antibody dilutions. Black lines represent the mean of the three antigens in each condition.

#### S1.2 Analytical assay performance

For assessment of analytical assay performance (intra- and inter-assay variability) as well as for quality control purposes, we generated two control samples, a positive control made from a pool of sera from PCR-confirmed COVID-19 cases and a negative control made from a pool of negative sera. These samples were tested in quadruplicates in the same and in separate 96-well plates at different days and MFI values were used to calculate the coefficient of variation (CV%) for intra- and inter-assay variability. Also, the ratio of the average MFI values of the positive against the negative control for the SARS-CoV-2 N, S1 and RBD antigens was calculated and used as an indicator for the proper performance of the assay procedure by the end-user. Assay performance was considered successful when a ratio of three or greater for all three antigens was achieved. CV% values for the three SARS-CoV-2 antigens were below 20% across all isotypes (Supplementary Table S1). The IgG assay performed better across all antigens, while the highest variability was observed in the antigens from the endemic coronaviruses in the total IgG/IgM/IgA assay.

**Supplementary Table S1.** Intra- and Inter-assay variability, measured as % coefficient of variation.

| Antibody Isotype | Variability | Sample | SARS-CoV-2 N | SARS-CoV-2 S1 | SARS-CoV-2 RBD | HKU1 S1 | NL63 S1 | 229E S1 | OC43 S1+S2 |
|---|---|---|---|---|---|---|---|---|---|
| Total (IgG/IgM/IgA) | Intra-assay | Positive serum | 8.2 | 8.2 | 8.2 | 20.3 | 23.5 | 20.7 | 12.2 |
| | | Negative serum | 15.2 | 13.8 | 8.3 | 18.0 | 19.9 | 16.2 | 13.7 |
| | Inter-assay | Positive serum | 4.3 | 4.3 | 4.1 | 11.5 | 13.7 | 11.0 | 5.6 |
| | | Negative serum | 15.8 | 16.5 | 8.2 | 28.9 | 30.6 | 24.4 | 21.3 |
| IgG | Intra-assay | Positive serum | 0.9 | 0.7 | 1.1 | 2.5 | 3.6 | 2.5 | 1.0 |
| | | Negative serum | 4.1 | 3.2 | 5.9 | 4.0 | 5.1 | 3.2 | 3.1 |
| | Inter-assay | Positive serum | 1.0 | 1.4 | 1.3 | 3.2 | 3.6 | 2.5 | 1.2 |
| | | Negative serum | 1.6 | 2.5 | 4.0 | 3.2 | 3.7 | 2.5 | 2.7 |
| IgA | Intra-assay | Positive serum | 7.4 | 8.2 | 8.6 | 12.8 | 13.7 | 12.3 | 9.8 |
| | | Negative serum | 4.6 | 6.7 | 4.7 | 5.5 | 6.2 | 5.1 | 4.1 |
| | Inter-assay | Positive serum | 10.0 | 11.3 | 11.3 | 17.5 | 17.7 | 16.1 | 13.2 |
| | | Negative serum | 4.9 | 7.2 | 5.0 | 7.1 | 8.0 | 6.5 | 4.6 |
| IgM | Intra-assay | Positive serum | 2.4 | 1.9 | 2.3 | 7.0 | 5.0 | 4.9 | 4.0 |
| | | Negative serum | 3.8 | 4.0 | 3.4 | 4.1 | 8.7 | 5.2 | 3.7 |
| | Inter-assay | Positive serum | 2.8 | 2.3 | 1.8 | 3.9 | 2.2 | 4.5 | 4.0 |
| | | Negative serum | 3.9 | 8.5 | 5.6 | 7.1 | 6.2 | 7.7 | 6.9 |

#### S1.3 Antibody responses to SARS-CoV-2 and endemic human coronaviruses in clinical samples

A total of 155 serum samples from 77 PCR-confirmed COVID-19 cases and 78 pre-epidemic individuals were screened for the existence of reactive antibodies against antigens of the five different coronaviruses (Supplementary Table S2). SARS-CoV-2 infection was deemed asymptomatic and mild needing no hospitalization in 8% and 60% of the cases, respectively, whereas 32% of the participants required hospitalization. Antibody detection was performed at a median of 46 days (range 13-87) post SARS-CoV-2 infection. Supplementary Figure S3 shows the reactivity of sera to each antigen for the different antibody isotypes. Antibody levels against the SARS-CoV-2 antigens N, S1 and RBD were significantly higher in the SARS-CoV-2 infected population as compared to samples from non-infected individuals ($p<0.001$, Supplementary Table S3); this finding was true for all isotypes. The analyzed blood samples from SARS-

CoV-2 tested positive donors also showed significantly high antibody titers against the HCoV-OC43 S1/S2 antigens in all isotypes and against the HCoV-HKU1 S1 antigen in IgG detection, possibly as a result of antibodies cross-reactivity against S proteins that are conserved between SARS-CoV-2, HCoV-HKU1 and HCoV-OC43 coronaviruses.

We further assessed the correlation of the normalized MFI values between antigens in both positive and negative cases (Supplementary Table 4). The SARS-CoV-2 S1 and RBD antigens-related data correlated strongly across all Ig isotypes in COVID-19 positive samples with Pearson's correlation coefficient (r) values > 0.96; a high correlation was also observed between N and S1 or RBD readouts in IgG and total antibody assays (r values between 0.72-0.82) in COVID-19 positive samples. Readouts between the N and S1 or RBD antigens correlated poorly in IgA and IgM detection. Results from negative samples showed no correlation across all antigen comparisons and Ig isotypes with the notable exception of the S1 and RBD antigens in IgM isotype (r=0.78).

**Supplementary Table S2.** List of samples used throughout this study.

| **Cohort type** | **Sample type** | **Sample source** | **No of samples** | **Demographics** | | |
|---|---|---|---|---|---|---|
| | | | | Median age (range) | Gender | Days post infection (range) |
| Clinical | Positive serum (SARS-CoV-2 PCR-confirmed cases) | Alexandra General Hospital, Athens | 60 | 49 (20-74) | 65% M 35% F | 46 (13-87) |
| | | University Hospital of Patras | 17 | | | |
| | Negative serum (banked samples from 2018-2019) | University Hospital of Patras | 78 | 57 (17-71) | 51% M 49% F | N/A |
| Population screen | Serum from blood donors | Multi-center/ University of Patras | 1225 | 42 (17-65) | 74% M 26% F | N/A |

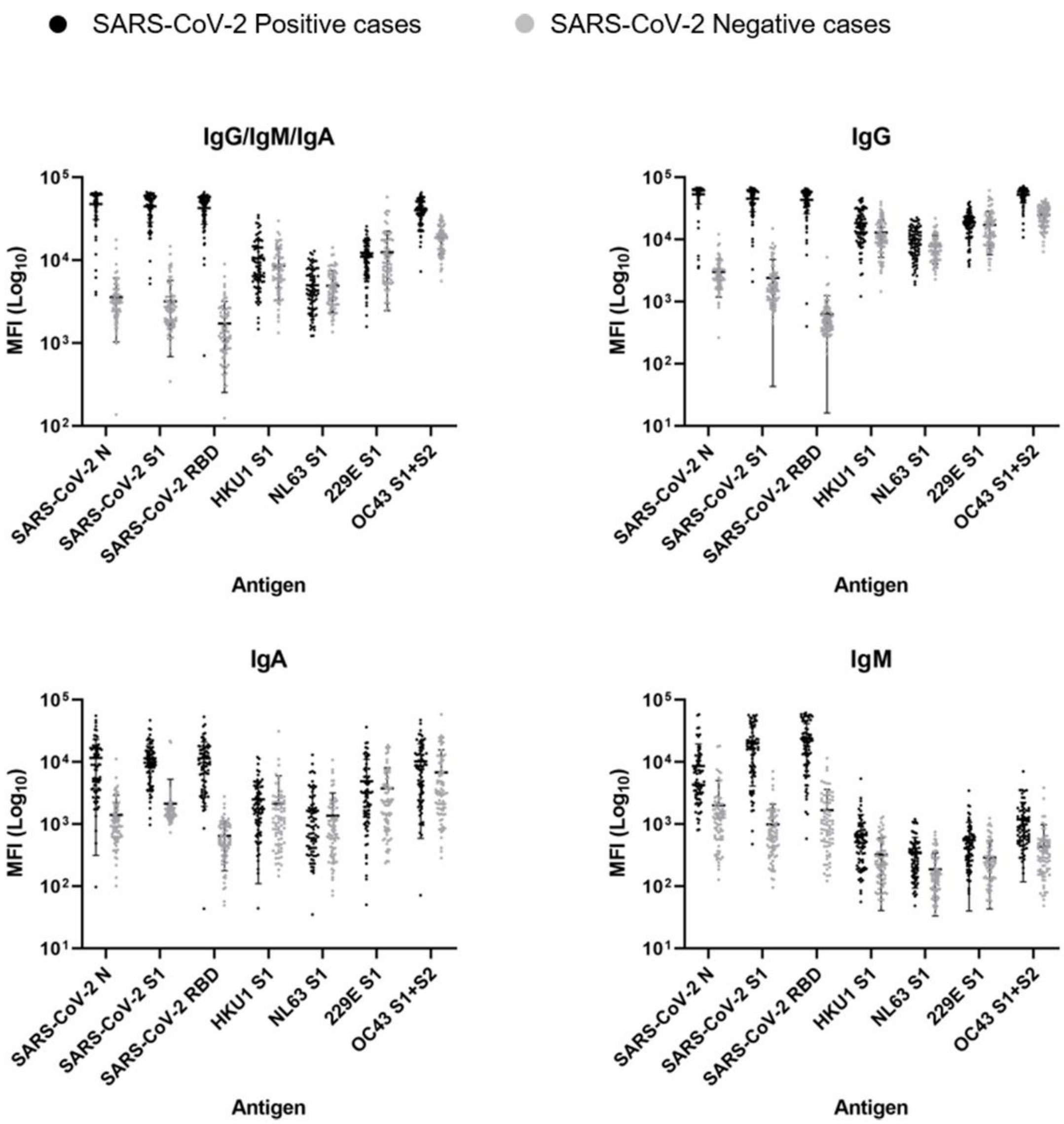


**Supplementary Figure S3.** Antibody responses against SARS-CoV-2 N, S1 and RBD antigens and 4 endemic coronaviruses antigens in serum samples from SARS-CoV-2 PCR-tested positive and negative (banked blood samples from 2018-2019) individuals. The Median Fluorescence Intensities (MFI, log10 scale) measured in positive and negative samples were plotted for each antigen. Graphs indicated the different antibody isotypes tested. Mean plus standard deviation (SD) error bars are presented.

**Supplementary Table S3.** Antibody responses to coronavirus antigens in SARS-CoV-2 positive and negative cases.

| | | SARS-CoV-2 Positive Cases | | SARS-CoV-2 Negative Cases | | |
|---|---|---|---|---|---|---|
| **Isotype** | **Antigen** | **Mean** | **SD** | **Mean** | **SD** | **P-Value** |
| Total (IgG/IgM/IgA) | SARS-CoV-2 N | 47598.8 | 16571.5 | 3571.9 | 2541.0 | <0.000001 |
| | SARS-CoV-2 S1 | 44681.6 | 15960.5 | 3188.2 | 2504.0 | <0.000001 |
| | SARS-CoV-2 RBD | 42692.9 | 15336.8 | 1721.6 | 1467.9 | <0.000001 |
| | HKU1 S1 | 10564.6 | 7393.9 | 8587.7 | 5259.1 | 0.185844 |

| | | | | | | |
|---|---|---|---|---|---|---|
| | NL63 S1 | 5011.9 | 2905.8 | 4918.9 | 2566.9 | 0.950362 |
| | 229E S1 | 11056.3 | 5040.5 | 12465.3 | 10001.0 | 0.345655 |
| | OC43 S1+S2 | 40071.6 | 12817.1 | 18794.9 | 6972.1 | <0.000001 |
| IgG | SARS-CoV-2 N | 53384.7 | 16219.0 | 3017.1 | 1824.5 | <0.000001 |
| | SARS-CoV-2 S1 | 45661.3 | 17638.3 | 2405.4 | 2362.3 | <0.000001 |
| | SARS-CoV-2 RBD | 43344.2 | 17129.8 | 628.3 | 612.2 | <0.000001 |
| | HKU1 S1 | 18405.4 | 10656.9 | 12968.2 | 7818.9 | 0.001355 |
| | NL63 S1 | 10008.0 | 5235.7 | 7828.3 | 3758.6 | 0.198085 |
| | 229E S1 | 18640.9 | 8172.8 | 17179.1 | 11478.5 | 0.387976 |
| | OC43 S1+S2 | 52576.7 | 13693.9 | 24740.7 | 9010.3 | <0.000001 |
| IgA | SARS-CoV-2 N | 11626.9 | 11311.3 | 1405.7 | 1518.3 | <0.000001 |
| | SARS-CoV-2 S1 | 11443.1 | 8100.1 | 2126.1 | 3151.4 | <0.000001 |
| | SARS-CoV-2 RBD | 11455.2 | 9523.9 | 646.2 | 469.3 | <0.000001 |
| | HKU1 S1 | 2497.7 | 2387.3 | 2143.3 | 3836.7 | 0.72526 |
| | NL63 S1 | 1644.2 | 2243.8 | 1367.7 | 1795.8 | 0.783876 |
| | 229E S1 | 4866.8 | 6111.9 | 3758.0 | 4360.1 | 0.271624 |
| | OC43 S1+S2 | 10162.1 | 9569.3 | 6781.4 | 8937.7 | 0.000826 |
| IgM | SARS-CoV-2 N | 8637.6 | 10910.7 | 1997.5 | 3019.2 | <0.000001 |
| | SARS-CoV-2 S1 | 20217.3 | 16141.8 | 986.4 | 1112.8 | <0.000001 |
| | SARS-CoV-2 RBD | 23718.1 | 17758.2 | 1679.1 | 1928.3 | <0.000001 |
| | HKU1 S1 | 675.0 | 741.6 | 321.3 | 280.7 | 0.756783 |
| | NL63 S1 | 351.6 | 258.9 | 187.2 | 154.0 | 0.885512 |
| | 229E S1 | 552.3 | 512.4 | 290.5 | 247.2 | 0.818666 |
| | OC43 S1+S2 | 1187.1 | 1068.8 | 439.5 | 534.0 | 0.512724 |

**Supplementary Table S4.** Correlation of normalized MFI values between SARS-CoV-2 antigens for the different antibody isotypes in positive and negative cases.

| **Isotype** | **Comparison** | **SARS-CoV-2 Positive Cases** | **SARS-CoV-2 Negative Cases** |
|---|---|---|---|
| Total (IgG/IgM/IgA) | N *vs* S1 | 0.82 | 0.29 |
| | N *vs* RBD | 0.81 | 0.32 |
| | S1 *vs* RBD | 0.98 | 0.23 |
| IgG | N *vs* S1 | 0.75 | 0.22 |

| | | | |
|---|---|---|---|
| | N *vs* RBD | 0.72 | 0.38 |
| | S1 *vs* RBD | 0.98 | 0.34 |
| | N *vs* S1 | 0.27 | 0.56 |
| IgA | N *vs* RBD | 0.24 | 0.21 |
| | S1 *vs* RBD | 0.96 | -0.03 |
| | N *vs* S1 | 0.39 | 0.37 |
| IgM | N *vs* RBD | 0.35 | 0.27 |
| | S1 *vs* RBD | 0.96 | 0.78 |

#### S1.4 Correlation of SARS-CoV-2 antibody responses to antibody levels from other infectious agents

For assessing interfering factors, we used results from 66 pre-epidemic fully characterized sera for the presence of antibodies against Cytomegalovirus (CMV), Epstein-Barr virus (EBV), Hepatitis A (HAV) and B (HBs) and Toxoplasma (Toxo) (Supplementary Table S5). CMV IgG levels positively correlated with SARS-CoV-2 S1 IgG and total (IgG/IgM/IgA) antibodies ($p<0.01$) but showed no significant correlation with antibody levels against N or RBD antigens (Supplementary Table S5). No correlation was observed between SARS-CoV-2 reactive antibodies and antibodies against the other infectious agents tested.

**Supplementary Table S5.** Correlation of SARS-CoV-2 antibody responses to antibody levels from other infectious agents.

| Isotype | Antigen | CMV | | EBV | | HAVAb | | Toxo | | HBs | |
|---|---|---|---|---|---|---|---|---|---|---|---|
| | | r | p-value | r | p-value | r | p-value | r | p-value | r | p-value |
| Total (IgG/IgM/IgA) | N | 0.24 | 0.062 | 0.29 | 0.026 | -0.23 | 0.080 | 0.12 | 0.359 | -0.05 | 0.719 |
| | S1 | 0.40 | 0.002 | 0.17 | 0.207 | 0.00 | 0.971 | 0.00 | 0.996 | -0.09 | 0.499 |
| | RBD | 0.07 | 0.608 | -0.05 | 0.701 | -0.18 | 0.179 | -0.04 | 0.754 | -0.03 | 0.840 |
| IgG | N | 0.19 | 0.156 | 0.32 | 0.013 | -0.13 | 0.335 | 0.12 | 0.357 | 0.00 | 0.982 |
| | S1 | 0.43 | 0.001 | 0.24 | 0.066 | 0.14 | 0.303 | 0.00 | 0.995 | -0.08 | 0.560 |
| | RBD | 0.06 | 0.666 | 0.22 | 0.101 | 0.05 | 0.699 | 0.00 | 0.973 | -0.08 | 0.546 |
| IgA | N | 0.33 | 0.011 | 0.11 | 0.402 | 0.09 | 0.491 | -0.03 | 0.849 | -0.07 | 0.578 |
| | S1 | 0.28 | 0.030 | 0.15 | 0.243 | 0.14 | 0.276 | -0.01 | 0.941 | -0.04 | 0.791 |
| | RBD | 0.17 | 0.192 | 0.11 | 0.421 | -0.12 | 0.368 | 0.08 | 0.573 | -0.11 | 0.415 |
| IgM | N | 0.21 | 0.102 | 0.14 | 0.280 | -0.18 | 0.173 | 0.01 | 0.928 | 0.03 | 0.794 |
| | S1 | 0.15 | 0.266 | -0.01 | 0.926 | -0.25 | 0.059 | -0.02 | 0.869 | -0.04 | 0.766 |
| | RBD | 0.11 | 0.410 | -0.07 | 0.618 | -0.20 | 0.137 | -0.05 | 0.686 | -0.01 | 0.946 |

#### S1.5 Receiver operating characteristic curve analysis

For each antibody ROC analysis is performed for the three (N, S1, RBD) antigens. The confidence intervals are calculated using Wilson's method. The AUC (area under the curve) of the ROC curves was calculated and used as a metric to evaluate the diagnostic performance of single antigen readouts (Supplementary Figure S4 & Supplementary Table S6).

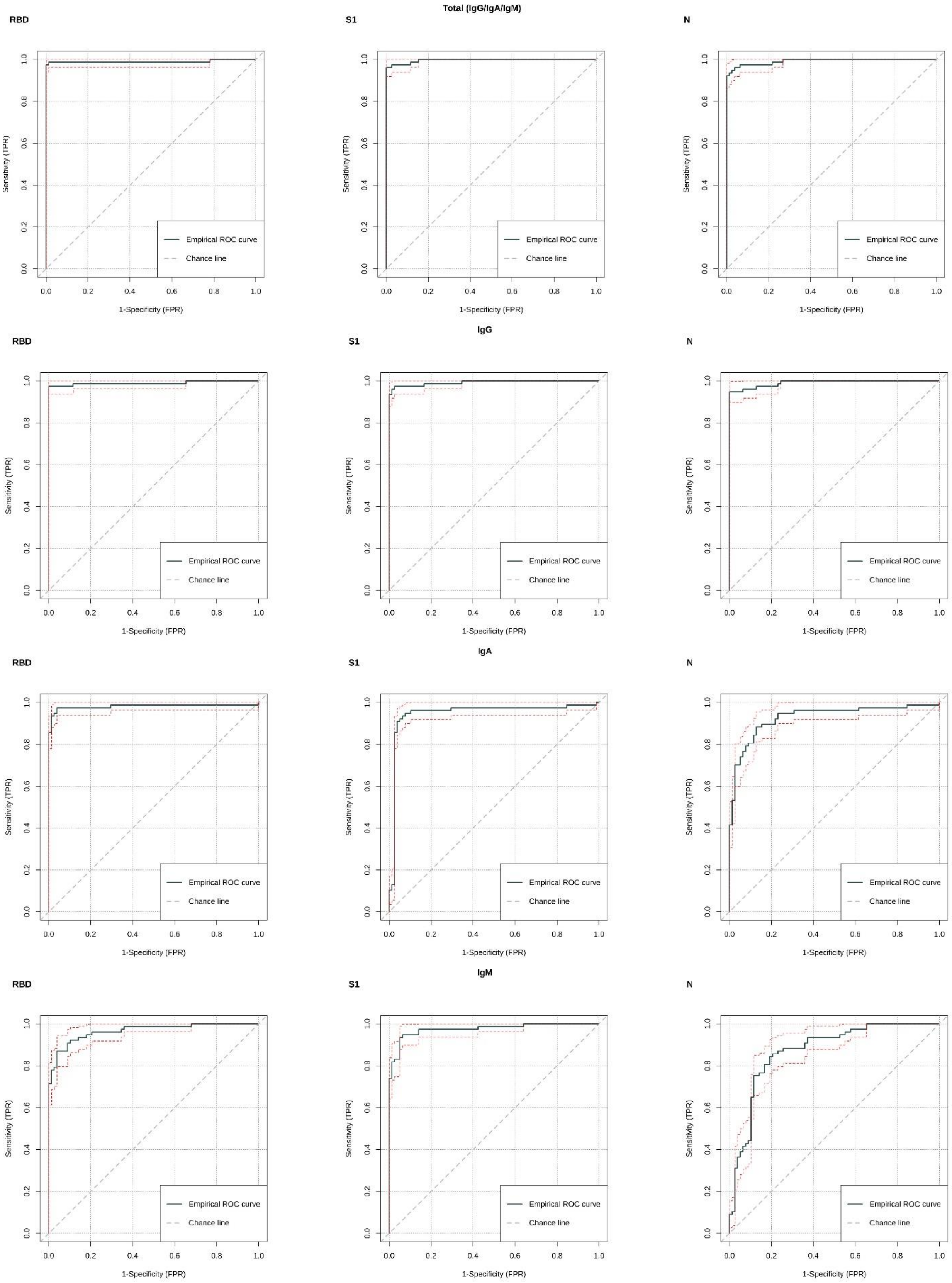
Total (IgG/IgA/IgM)
RBD
S1
N
IgG
RBD
S1
N
IgA
RBD
S1
N
IgM
RBD
S1
N
Sensitivity (TPR)
1-Specificity (FPR)
Empirical ROC curve
Chance line

**Supplementary Figure S4.** ROC curves for the 3 antigens and different antibody isotypes. Confidence intervals are demonstrated with red dotted lines.

**Supplementary Table S6.** Diagnostic accuracy and AUC of SARS-CoV-2 individual antigens.

| **Total (IgG/IgM/IgA)** | | | **IgG** | | **IgA** | | **IgM** | |
|---|---|---|---|---|---|---|---|---|
| **Antigen** | **Accuracy** [95% CI] | **AUC** [95% CI] | **Accuracy** [95% CI] | **AUC** [95% CI] | **Accuracy** [95% CI] | **AUC** [95% CI] | **Accuracy** [95% CI] | **AUC** [95% CI] |
| N | 0.961 [0.918,0.982] | 0.992 [0.973, 1.000] | 0.961 [0.918,0.982] | 0.991 [0.980, 1.000] | 0.755 [0.681,0.816] | 0.928 [0.885, 0.971] | 0.600 [0.521,0.674] | 0.872 [0.815, 0.929] |
| S1 | 0.968 [0.927,0.986] | **0.996** [0.986, 1.000] | 0.968 [0.927,0.986] | **0.993** [0.979, 1.000] | 0.677 [0.600,0.746] | 0.948 [0.911, 0.984] | **0.903** [0.845,0.940] | **0.975** [0.950, 1.000] |
| RBD | **0.981** [0.945,0.993] | 0.990 [0.973, 1.000] | **0.981** [0.945,0.993] | 0.990 [0.974, 1.000] | **0.955** [0.910,0.978] | **0.981** [0.959, 1.000] | 0.884 [0.824,0.925] | 0.966 [0.937, 0.996] |

#### S1.6 Seroprevalence and agreement rates of single and multi-antigen rules in the population screening

Seroprevalence rates calculated using the best performing multi-antigen rules for different cut-off values are presented in Supplementary Figure S5. The agreement rates between all multi-antigen rules and between rules and commercial tests are presented in Supplementary Figures S6 & S7.

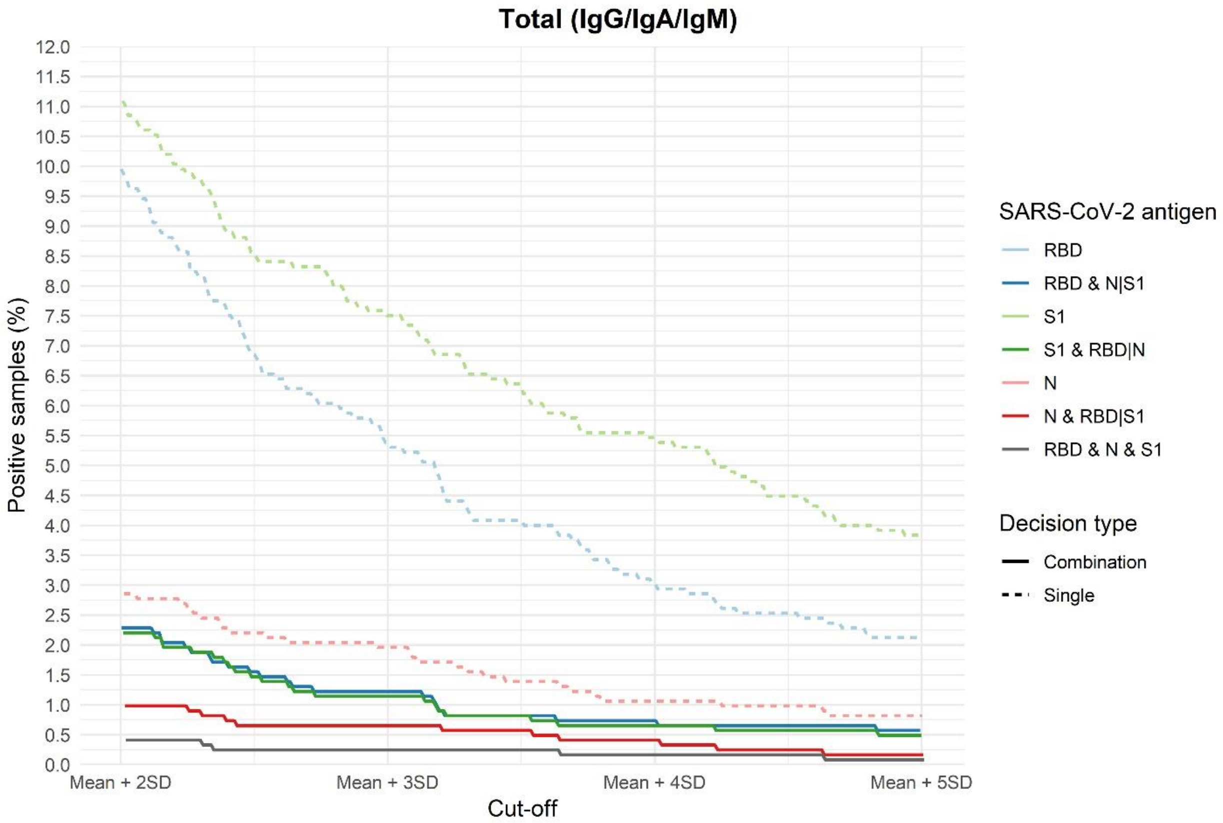


**Supplementary Figure S5.** Percentage of asymptomatic individuals (n=1,255) positive for total (IgG/IgA/IgM) SARS-CoV-2 antibodies using single antigen readouts and multi-antigen rules. Seroprevalence is calculated using different cut-off values from the distribution of negative samples used in the diagnostic performance analysis.

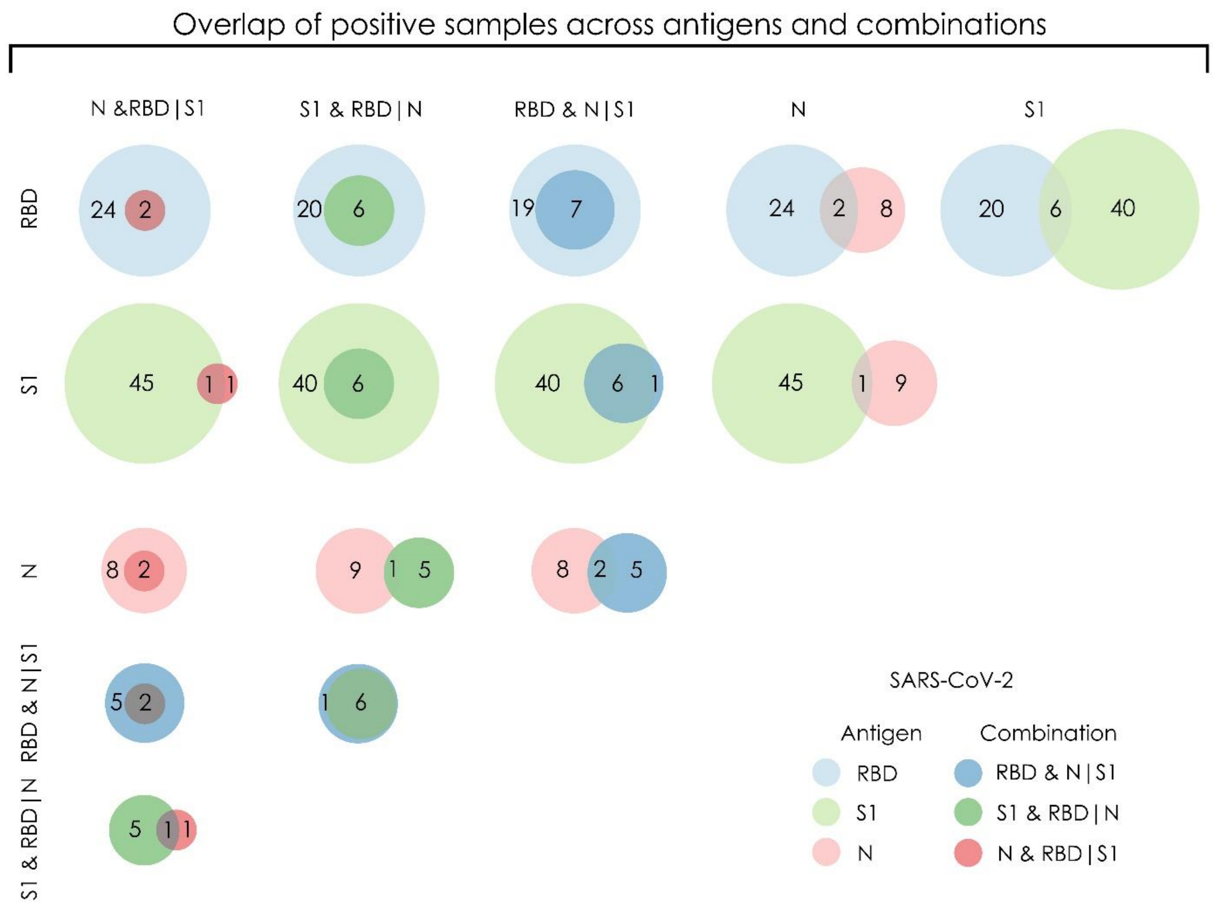


**Supplementary Figure S6.** Consensus between positive samples for total (IgG/IgA/IgM) antibodies using N, S1, RBD and respective multi-antigen rules in the population screen.

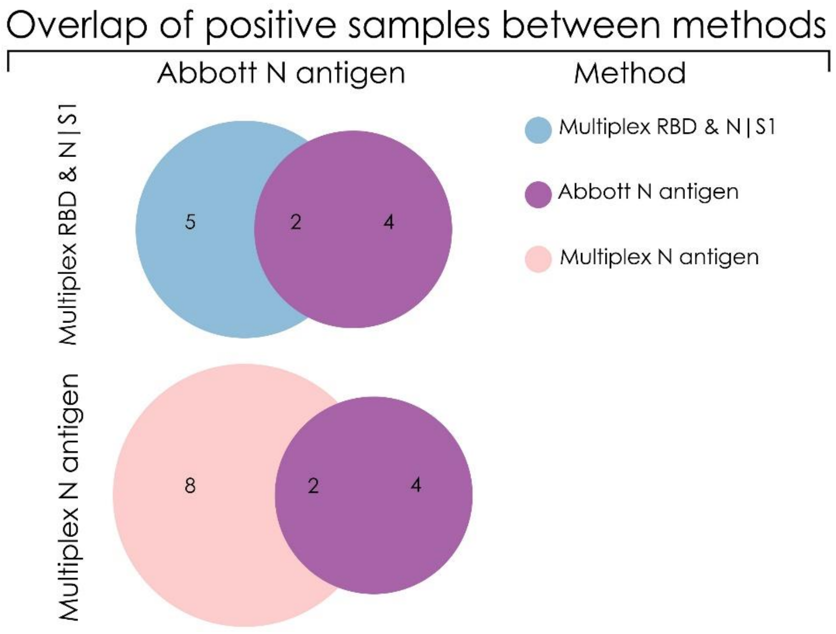

**Supplementary Figure S7.** Consensus between positive samples for total (IgG/IgA/IgM) antibodies against N, S1 and RBD and for IgG antibodies against N antigen (Abbott) in the population screen.

### S.2 Supplementary methods

#### S.2.1. Multiplex immunoassay development

A magnetic bead-based immunoassay was developed using the xMAP Luminex technology against SARS-CoV-2 antigens N, S1 and RBD. One antigen from each one of the four endemic coronaviruses was also included in the assay. Specifically, the S1 subunit of HCoV-HKU1, HCoV-229E and HCoV-NL63 and the S1+S2 subunits from HCoV-OC43 were used. The SARS-CoV-2 N and S1 antigens were purchased from the Native Antigen Company (Kidlington, UK). All other antigens were from Sino Biological Europe GmbH (Eschborn, Germany). Each antigen was covalently coupled to a distinct magnetic bead region (Luminex Corp, Austin, Texas) by carbodiimide coupling at a ratio of 15 µg per 5 million beads. Coupling efficiency was confirmed by incubation of 5,000 beads from each coupled region with a phycoerythrin-conjugated anti-6x HisTag antibody (Abcam, Cambridge, UK) at a concentration of 32 µg/mL for 15 min at room temperature. Coupled beads were mixed to a final concentration of 50 beads/µL and stored in PBS supplemented with 1% bovine serum albumin, 0.02% Tween-20 and 0.05% sodium azide at 4oC until use. For analysis of serum samples, 25 µL of the bead mix (corresponding to 1,250 beads per antigen) were added to each well of a 96-well plate, washed twice with 100 µL Assay Buffer (PBS supplemented with 1% BSA and 0.05% sodium azide) and incubated with 50 µL of serum diluted in LowCross-Buffer® (CANDOR Bioscience GmbH, Wangen, Germany) for 2 hrs at room temperature in a plate shaker (900 rpm). A serum dilution of 1/400 was used for testing all immunoglobulin types except for IgA that was assayed at a 1:100 serum dilution was used. Unbound material was removed by two washes with 100 µL assay buffer and beads were incubated with 20 µL of biotinylated anti-human immunoglobulin antibodies (Jackson ImmunoResearch Europe Ltd, Ely, UK) for 1 hr at room temperature in a plate shaker (900 rpm). Antibodies were diluted in assay buffer at 1:1,600 for IgG/IgM/IgA, 1:800 for IgG, 1:3,200 for IgA and 1:800 for IgM. Beads were washed twice with 100 µL assay buffer and incubated with streptavidin R-phycoerythrin (Jackson ImmunoResearch Europe Ltd, Ely, UK) diluted 1:100 in assay buffer for 15 min at room temperature in a plate shaker (900 rpm). Beads were washed again as before, reconstituted in 130 µL assay buffer, and measured in a FLEXMAP 3D instrument (Luminex Corp, Austin, Texas). Instrument settings included standard PMT, 100 µL sample volume, a bead count of 50 beads per antigen and doublet discrimination gate set at 3,000-20,000.

#### S.2.2 Clinical and donor samples

The list of all samples utilized in this study is presented in Supplementary Table S2.

#### S.2.3 Assay validation

Assay validation was performed in two separate subsets of matched clinical samples against two widely used, commercially available SARS-CoV-2 antibody tests developed by Euroimmun (Euroimmun Medizinische Labordiagnostika AG, Lubeck, Germany) and Abbott (Abbott Diagnostics, Illinois, USA), which detect IgG antibodies against S1 and N respectively (Supplementary Table S7).

**Supplementary Table S7.** Commercial SARS-CoV-2 serological assays and samples used for assay validation.

| Commercial Assay | Antigen Tested | Total No of Samples | Sample Type | Sample Source |
|---|---|---|---|---|

| | | | | |
|---|---|---|---|---|
| Euroimmun SARS-CoV-2 ELISA IgG | S1 | 60 | Positive Serum (SARS-CoV-2 PCR-tested positive donors)* | Alexandra General Hospital, Athens |
| Abbott<br><br>SARS-CoV-2 IgG | N | 31 | Positive Serum (SARS-CoV-2 PCR-tested positive donors)*<br><br>(n=12) | University Hospital of Patras |
| | | | Serum from SARS CoV-2 positive or negative individuals (n=19) | University Hospital of Patras |
